# Decoding the Best Automated Segmentation Tools for Vascular White Matter Hyperintensities in the Aging Brain: A Clinician’s Guide to Precision and Purpose

**DOI:** 10.1101/2023.03.30.23287946

**Authors:** Lucia Torres-Simon, Alberto del Cerro-León, Miguel Yus, Ricardo Bruña, Lidia Gil-Martinez, Alberto Marcos Dolado, Fernando Maestú, Juan Arrazola-Garcia, Pablo Cuesta

## Abstract

Cerebrovascular damage from small vessel disease (SVD) occurs in healthy and pathological aging. SVD markers, such as white matter hyperintensities (WMH), are commonly found in individuals over 60 and increase in prevalence with age. WMHs are detectable on standard MRI by adhering to the STRIVE criteria. Currently, visual assessment scales are used in clinical and research scenarios but is time-consuming and has rater variability, limiting its practicality. Addressing this issue, our study aimed to determine the most precise WMH segmentation software, offering insights into methodology and usability to balance clinical precision with practical application. This study employed a dataset comprising T1, FLAIR, and DWI images from 300 cognitively healthy older adults. WMHs in this cohort were evaluated using four automated neuroimaging tools: Lesion Prediction Algorithm (LPA) and Lesion Growth Algorithm (LGA) from Lesion Segmentation Tool (LST), Sequence Adaptive Multimodal Segmentation (SAMSEG), and Brain Intensity Abnormalities Classification Algorithm (BIANCA). Additionally, clinicians manually segmented WMHs in a subsample of 45 participants to establish a gold standard. The study assessed correlations with the Fazekas scale, algorithm performance, and the influence of WMH volume on reliability. Results indicated that supervised algorithms were superior, particularly in detecting small WMHs, and can improve their consistency when used in parallel with unsupervised tools. The research also proposed a biomarker for moderate vascular damage, derived from the top 95th percentile of WMH volume in healthy individuals aged 50 to 60. This biomarker effectively differentiated subgroups within the cohort, correlating with variations in brain structure and behavior.

## 1 Introduction

White matter hyperintensities (WMH) can be observed in magnetic resonance image (MRI) among older adults, with or without dementia [1], [2], being small vessel disease (SVD) the most common underlying cause [3]. The significance of white matter hyperintensities (WMH) is well-documented, with strong associations to dementia, particularly vascular cognitive impairment (VCI), and to the lifetime risk of stroke [4], [5]. Research indicates that WMH volume correlates with cognitive decline, disability progression, and overall prognosis [6], [7]. These effects are evident in both cognitively healthy seniors [8], [9] and those with dementia [10], [11], impacting attention and executive function. Moreover, WMHs have implications for mental health, contributing to conditions like depression and anxiety [12], [13]. Pathologically, WMHs are linked to broader brain changes, including amyloid and tau deposits [14], [15], grey matter atrophy [14], [16], ventricular enlargement [17], and disruptions in structural [18] and functional [19], [20] connectivity. Consequently, the careful monitoring of WMHs is crucial for the early identification and management of individuals at risk, underscoring its importance in research and clinical practice.

Per STRIVE guidelines, WMH are identified as diffuse, bright areas (hence, “hyperintense”) on T2-weighted or FLAIR MRI scans, crucial for diagnosing and tracking white matter deterioration [21], [22]. Nowadays, visual assessment is the most used method, being the Fazekas scale [23] and the age-related white matter changes scale (ARWMC) (Wahlund et al., 2001) the most recommended. Nevertheless, these scales lack the precision needed for quantifying subtle progressions in WMH. Comprehensive MRI analysis of WMH— including number, volume, and distribution—can inform on disease etiology and prognosis [25], [26]. Moreover, quantifiable WMH biomarkers could provide objective thresholds for clinical decisions, such as treatment eligibility [27], [28]. In this context, manual segmentation is the most precise protocol for quantifying WMH [29], [30] but has high inter- and intra-rater variability and is too time-consuming and for practical use. Consequently, there’s a growing demand for automatic segmentation tools, which are either supervised, requiring a pre-labeled training set, or unsupervised, not depending on a gold standard [31]. These tools, primarily designed for studying multiple sclerosis [32], [33], must be evaluated for their effectiveness in identifying WMH related to vascular changes in aging [25]. Comprehensive reviews [25], [31] have previously analyzed such automated methods, but the diversity of tools and methodologies yields inconclusive results that are not yet fully applicable in clinical settings. In addition, consideration of WMH load on the performance of the segmentation software is scarce but crucial in scenarios where the amount of injury ranges from low to moderate proportions like ageing.

Our study aims to evaluate widely used neuroscience software packages (SPM12, Freesurfer & FSL) to identify the best algorithm for automatic WMH volume measurement, tailored to user needs. We assessed four algorithms: 1) SPM12’s lesion segmentation tool - lesion growth algorithm (LST-LGA); 2) SPM12’s lesion segmentation tool - Lesion prediction algorithm (LST-LPA); 3) FreeSurfer’s sequence adaptive multimodal segmentation (SAMSEG); and 4) FSL’s brain intensity abnormality classification algorithm (BIANCA) in 45 cognitively healthy seniors with varying WMH levels, using expert manual segmentation as the gold standard. Correlations between algorithmic and manual WMH total volumes, as well as the Fazekas scale, were examined, alongside algorithm performance metrics. The top algorithms were then tested on a larger database of 300 individuals to determine their clinical and research utility, including identifying a WMH volume threshold with potential clinical significance.

## 2 Materials & Methods

### 2.1 Participants

The participants in the study were sourced from an established database designed for dementia research and detection, compiled from 2010 to 2018 through a collaborative effort with three clinical centers in Madrid: The Neurology Department in “Hospital Universitario Clínico San Carlos”, the “Centro de Prevención del Deterioro Cognitivo”, and the “Centro de Mayores del Distrito de Chamartín”. We initially recruited 520 cognitively healthy Spanish speakers. Each participant completed a thorough set of neuropsychological evaluations, genetic profiling, and neuroimaging assessments, which included structural MRI scans: T1, FLAIR, and diffusion-weighted imaging (DWI) (see **supplementary material 1** for a detailed description of the neuropsychological evaluation). Exclusion criteria included a history of psychiatric or neurological disease and psychoactive or chronic medication use. We also conducted tests (class II evidence level) to rule out other causes of cognitive decline such as B12 deficiency, diabetes mellitus, thyroid problems or infections. After informed consent and ethical clearance, participants underwent MRI scans. We excluded (number in brackets) individuals based on age below 50 (26), Mini Mental State Examination [34] score under 26 (22), brain abnormalities other than WMH (24), inadequate MRI data (100), and excessively high WMH volumes (4). The final sample comprised 300 individuals (Age = 66 ± 8; 68% females) that were employed in the following analyses (**Figure 1**).

**Figure 1.**
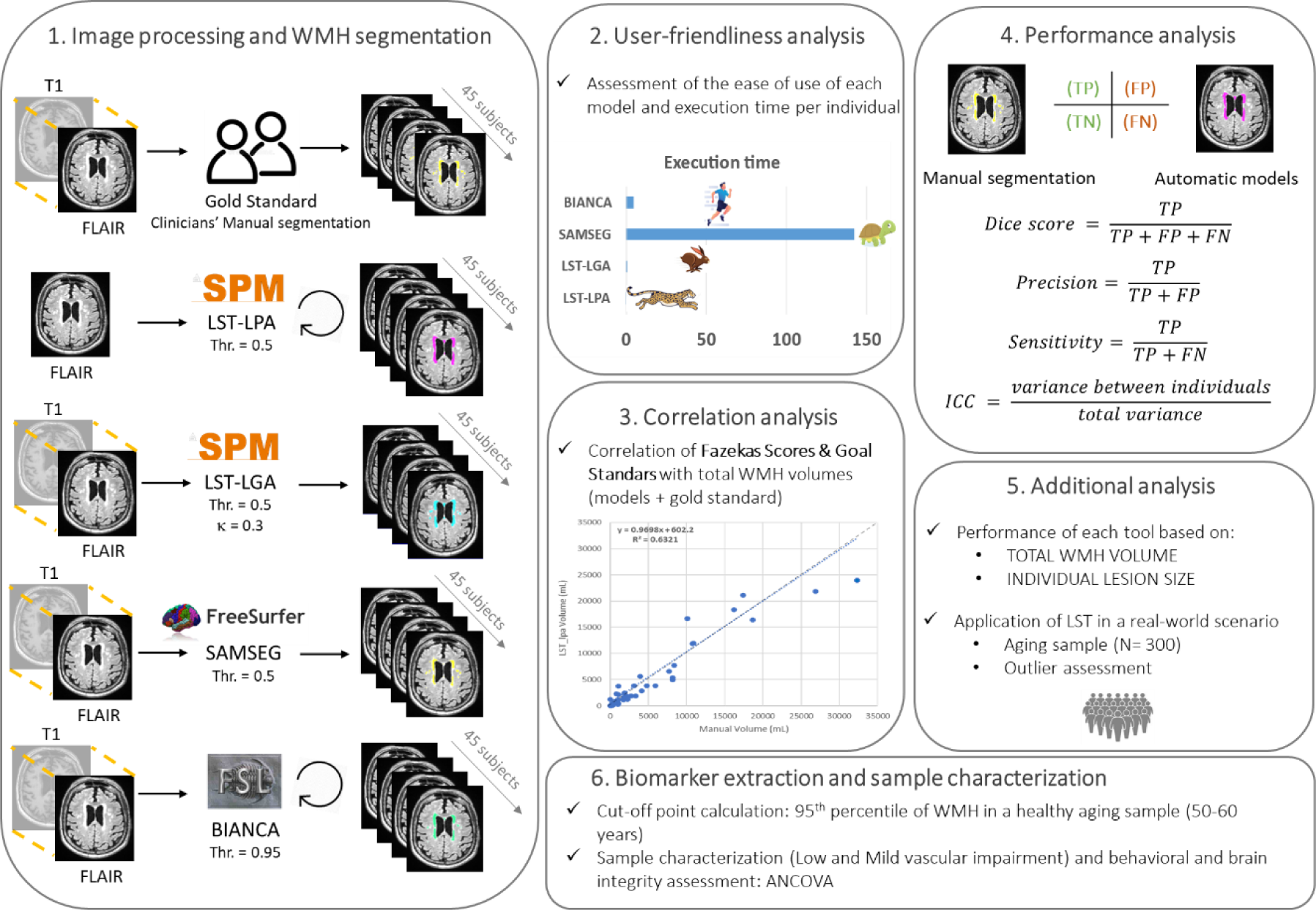
Methodological pipeline. From left to right: **1:** 3D T1 weighted and 3D FLAIR images co-registration, when required. Optimization of each algorithm performance and computation of the WMH masks of each participant. The circular arrows represent the algorithms that needs a training set. **2:** Assessment of User-friendliness by means of 3 expert neuroscientists. **3:** Battery of comparison based on performance and clinical relevance. **4:** comparison of the WMH masks computed by each algorithm. **5:** Evaluation of the models’ performance based on different WMH volumes, when evaluating at the individual lesion level and when testing the algorithm in a large dataset **6**: Biomarkers extraction and evaluation of behavioral and structural integrity differences in function of vascular damage.

### 2.2 MRI acquisition

Each participant was subjected to an MRI assessment, which included a 3D T1-weighted, a 3D T2-weighted FLAIR image, and a DWI, obtained with a General Electric 1.5 Tesla magnetic resonance scanner. The T1 and FLAIR images were acquired using a high-resolution antenna and a homogenization PURE filter (Fast Spoiled Gradient Echo sequence). The parameters for the T1 image were repetition time (TR) of 11.2 ms, echo time (TE) of 4.2 ms, inversion time (TI) of 450 ms, a field of view (FOV) of 25 cm, a flip angle (FA) = 12°, obtaining 252 coronal slices (in-plain resolution of 256×256) with a voxel size of 0.98 x 0.98 x 1 mm^3^, and an acquisition time ≃ 8:00 min. The T2-weighted 3D FLAIR images were obtained using a TR of 7000 ms, a TE of 101 ms, a TI of 2112 ms, a FOV of 24 cm, obtaining 252 sagittal z-axis interpolated slices (in-plain resolution of 256×112) with a voxel size of 0.94 x 0.94 x 1.6 mm^3^ and an acquisition time ≃ 4:57 min. Finally, the DWI images were acquired using a single shot echo planar imaging sequence with a TE of 96.1 ms, a TR of 12ms; a NEX 3 for increasing the signal-to-noise ratio, a FOV of 30.7 cm (in pain resolution of 128 x 128), a 4 mm slice thickness, yielding an isotropic voxel of 2.4 mm^3^. In the DWI sequence, on image had no diffusion sensitization (i.e., T2-weighted b0 images); and 25 were DWI (b = 900 s/mm2).

### 2.3 MRI clinical assessment: Fazekas Scores and WMH manual segmentation

Each participant’s FLAIR images were aligned with their T1 images for a unified anatomical reference. The radiology team at "Hospital Universitario Clínico San Carlos" manually outlined the WMH on the FLAIR images, consulting the T1 images as needed. They used FSLeyes (McCarthy, 2018) on a standard monitor, to ensured uniformity in segmentations. Discrepancies were resolved by two senior radiologists to set a consistent gold standard. The segmented WMH were then grouped into a 3D map of individual lesions per subject, and a Fazekas score was assigned to reflect WMH severity. See **Figure 2** for a visual indication of the Fazekas scores.

**Figure 2.**
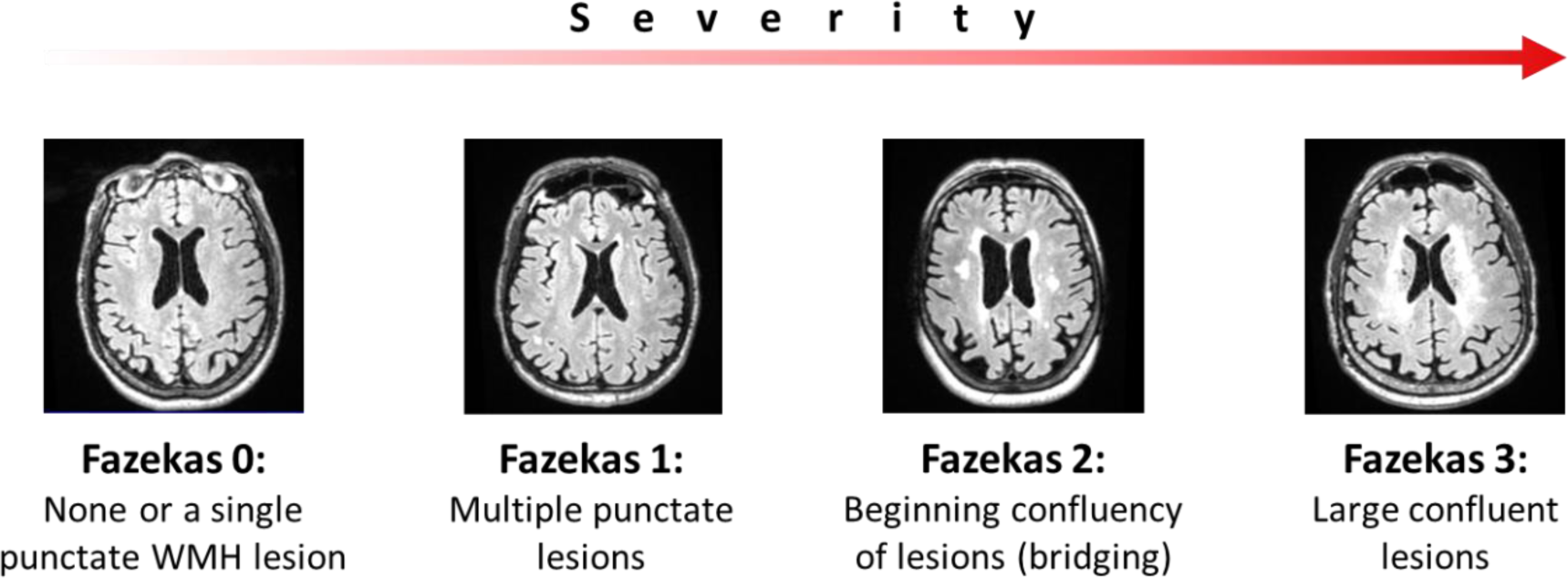
Explanation of the Fazekas scale, from left to right the different degrees of severity are detailed from lowest to highest.

### 2.4 Automatic WMH segmentation

WMH quantification from FLAIR/T1 images was conducted using four algorithms (**Figure 1, panel 1**), illustrated in Figure 2, panel 1. Within SPM12, we utilized the Lesion Segmentation Tool’s "lesion growth algorithm" (LST-LGA) and "lesion prediction algorithm" (LST-LPA), representing unsupervised and supervised methods respectively. From FSL, the "Brain intensity abnormality classification algorithm" (BIANCA), a supervised method, was employed. Additionally, from FreeSurfer, the unsupervised "Sequence adaptive multimodal segmentation" (SAMSEG) was applied. For an in-depth explanation of each method, please refer to the **Supplementary Material 2**. Additionally, we considered FreeSurfer’s T1-based WMH segmentation for being the most accessible in case of having only a T1 image.

### 2.5 Grey and white matter integrity

The T1 images were processed using the FreeSurfer software (version 6.1.0) for automated cortical and subcortical segmentation. [35]. The measures that were included in further analyses were total volumes (mm^3^) of gray matter, total white matter, lateral ventricles, and hippocampus; and the cortical thickness (mm). The volumes of bilateral structures were collapsed to obtain a single measure per region.

DWI images were processed with AutoPtx (https://fsl.fmrib.ox.ac.uk/fsl/fslwiki/AutoPtx) for probabilistic tractography, as outlined by [36]. The procedure extracted the mean fractional anisotropy (FA) for each tract, combining bilateral tracts into single measures. For a detailed description see **Supplementary materials 3**.

### 2.6 Algorithm performance analysis

Out of the 300 participants, we randomly selected 45 based on their clinical reports to represent a spectrum of WMH loads: 15 with no alterations in their MRI, 15 with subclinical WMH, and 15 with WMH presumably associated with cerebrovascular disease (CBVD). This selection ensured a diverse range of WMH loads for the study. The demographic and clinical data at baseline evaluation for these participants are described in **Table 1**.

**Table 1.** Demographic and clinical data. Values are presented as mean ± standard deviation. Sex (female/male), age (in years), education (expressed in years of education). MMSE: Mini Mental State Examination. WMH volume (in mm^3^) corresponded with those obtained with the radiologist’s manual segmentation. Demographics are displayed for the whole sample and for each subsample of interest.

|  | N | Age | Sex | Education | MMSE | Fazekas | Volume WMH |
| --- | --- | --- | --- | --- | --- | --- | --- |
| **Whole sample** | 45 | 71 ± 5 | 30/15 | 14 ± 6 | 29 ± 1 | 0.9 ± 0.8 | (5 ± 7) · 10<sup>3</sup> |
| **No WMH** | 15 | 69 ± 5 | 11/4 | 12 ± 5 | 28 ± 1 | 0.2 ± 0.4 | (1 ± 2) · 10<sup>3</sup> |
| **Subclinical WMH** | 15 | 71 ± 5 | 11/4 | 13 ± 6 | 29 ± 1 | 0.7 ± 0.5 | (2 ± 1) · 10<sup>3</sup> |
| **CBVD WMH** | 15 | 72 ± 4 | 8/7 | 15 ± 6 | 28 ± 2 | 1.8 ± 0.6 | (13 ± 8) · 10<sup>3</sup> |

#### User-friendliness

To address the user-friendliness (**Supplementary Material 4**) of each algorithm, three neuroimaging experts (authors L.T-S, A.C-L and P.C.) used each of the tools and indicated whether they had encountered any of the following difficulties during their execution: 1) *installation packages* (if the algorithm is included or it requires one or more additional packages); 2) *file preparation* (making reference to the images pre-processing required before the segmentation); 3) *brain extraction* (if it is required before segmentation process); 4) *training set* (if it is required for the algorithms and when it is, if this set is included or the user should provide its own); 5) *interface* (if there is an user-friendly interface or the user should work in the terminal environment); 6) *RAM requirements* (amount of available RAM required to run the algorithms); and 7) *execution time* (which was calculated for each algorithm as the corresponding time (in minutes) expended for the computation of the WMH masks, without considering the images or dataset preparations and/or preprocessing).

#### Correlation analysis

We conducted Spearman correlation analyses to compare WMH total volumes from automatic algorithms and the gold standard with Fazekas scores, assessing the accuracy of segmentation methods against the widely used clinical visual rating scale. Additionally, we compared total lesion volumes from each algorithm to those from the gold standard (**Figure 1, panel 3**).

#### Mean performance

Segmentation quality was assessed by comparing algorithm-generated binary masks with the gold standard (**Figure 1, panel 4**), accounting for true positives (TP), true negatives (TN), false positives (FP) and false negatives (FN). Consequently, we calculated 4 reliability metrics: 1) interclass correlation coefficient, reporting consistency and agreement between metrics; 2) Dice score, reporting the degree of similarity between automatic segmentation and the gold standard mask [37]; 3) precision, indicating the percentage of the segmented lesion that corresponds to an real lesion; and 4) sensitivity, indicating the percentage of the lesion present in the gold standard and captured by the automatic tool. The mathematical expression per each measure is depicted in the **Figure 1, panel 4**. The scores 2-4 were obtained per participant, obtaining three series of 45 values. Performance differences between tools were tested using paired t-tests and Hedges-corrected Cohen’s D

#### Performance based on WMH load

Regression analyses determined the impact of lesion volume on precision, sensitivity, and Dice scores for each segmentation tool (**Figure 1, panel 5**). To evaluate the tools’ performance relative to individual lesion size, all manual segmentations of the gold standard were clustered using a labeling algorithm based on elements connected in 26 directions in a binary matrix Only lesions comprising at least 4 voxels were included. Then, regardless of the participant, lesions were pooled together to create a data set of 1960 lesions. The sensitivity was measured in each lesion separately, and the results were sorted by volume into 10 groups of 196 lesions, ascending order of size. For each group of lesions, One-tailed t-tests evaluated if tool sensitivity significantly exceeded 0.5 for each volume group.

#### Outliers’ detection

We tested the LST-LGA and LST-LPA algorithms’ performance in a real-world context by evaluating total WMH across the entire sample (n = 300) (**Figure 1, panel 5**). For each participant, we computed the absolute difference in WMH volumes yielded by the two algorithms. We then determined the mean and standard deviation of these discrepancies for the sample, setting an outlier threshold at three standard deviations above the mean. An expert in neuroimaging (author A.C.L) manually reviewed any cases with differences exceeding this threshold.

### 2.7 Biomarker extraction

To establish a threshold at which WMH volume in the brain may have clinical significance, we calculated a specificity-based cutoff following the guidelines for defining imaging cut points [38]. Consistent with the study’s recommendations, only clinically normal individuals were included to identify potential biomarkers. We utilized WMH volumes from automatic segmentations of participants aged 50 to 60 years without evidence of brain damage, including vascular disease. This resulted in a subsample of 82 individuals (mean age 55 ± 3 years). For these subjects, the cutoff was determined to be the 95th percentile of the WMH volume distribution.

#### Sample characterization according to the cerebrovascular damage (WMH)

Based on this cutoff point, we divided the participants into two subgroups: ow vascular load (Low-VL) below the WMH threshold and mild vascular load (Mild-VL) exceeded it (**Figure 1, panel 6**). After establishing both groups, we assessed differences in demographic, cognitive, and anatomical variables using ANCOVA tests between groups, adjusting for age and total brain volume as covariates. We then adjusted the resulting p-values for multiple comparisons using a false discovery rate (FDR) of 0.05 [39].

### 2.8 Data availability

The data that support the findings of this study are available from the corresponding author upon reasonable request. All the algorithms used in the present paper are reported in the ‘Materials and methods’ section.

## 3 Results

The performance of each WMH segmentation algorithm was evaluated using the procedure outlined in the methods section (**Figure 1**). This sequence was designed to guide researchers through the necessary steps to master the methods, comprehend the clinical implications of the results, and apply them to a population of older adults.

### 3.1. Correlation analysis

We assessed the correlation between total WMH volumes from each tool (and the gold standard) and clinical Fazekas scores using Spearman correlations. The gold standard had the strongest association (R^2^ = 0.71) with Fazekas scores. All tools showed significant correlations (R² > 0.61), with BIANCA demonstrating the highest variability explained (R² = 0.64) among automated methods (see **Supplementary material 5**). Correlation analyses revealed all tools underestimated WMH volume compared to the gold standard, with LST-LPA being closest (slope of 0.97). LST-LGA and SAMSEG had slopes of 0.89 and 0.86, while BIANCA was furthest with a slope of 0.73 (graphical representations of these correlations can be found in **Supplementary material 6**). Regarding model fitting, BIANCA had the best linear regression fit (R² = 0.91), followed by LST-LGA (R² = 0.89), SAMSEG (R² = 0.86), and LST-LPA (R² = 0.62). The results of FreeSurfer package are available in **Supplementary material 7A**.

### 3.2. Performance analysis: mean scores

Each tool’s performance was gauged by comparing WMH brain masks to the gold standard, using interclass correlation coefficient (ICC), precision, sensitivity, and Dice scores (**Figure 3**). We also used paired samples t-tests to check for significant method differences in sensitivity, precision, and Dice scores.

**Figure 3.**
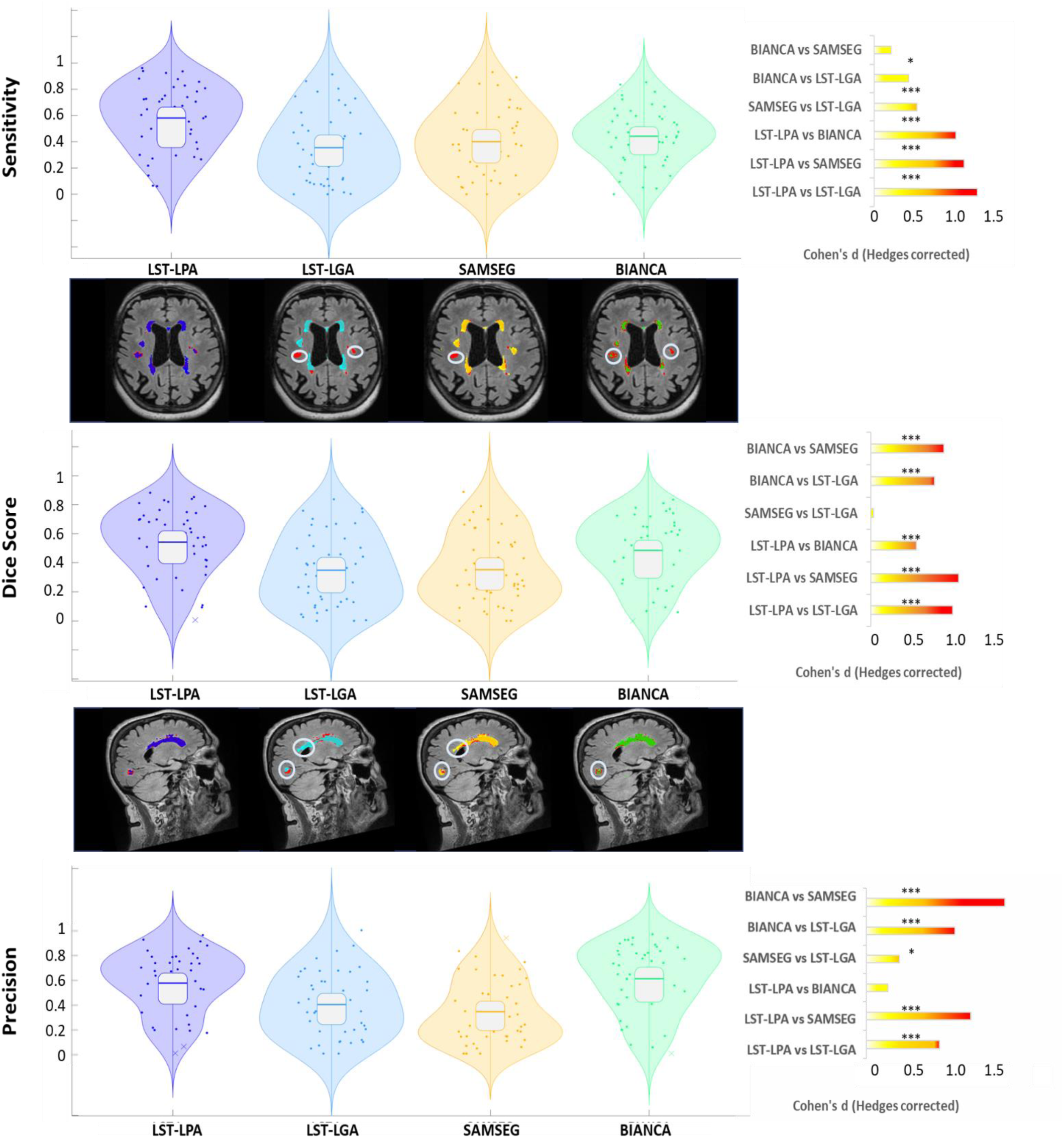
Lesion segmentation performance in terms of precision, sensitivity, and Dice score for the proposed methods (from left to right: LST-LPA, LST-LGA, SAMSEG and BIANCA). Violin plots depict the distribution of the values obtained in each tool LST-LPA (dark blue), LST-LGA (light blue), SAMSEG (yellow) and BIANCA (green). Within each violin plot, boxplots indicate the quartile positions. MRI images provide examples of segmentations performed by the various tools. Lesions identified by the gold standard are shown in red; those by LST-LPA in dark blue, LST-LGA in light blue, SAMSEG in yellow, and BIANCA in green. Gray circles on the MRI images highlight the principal differences among the tools’ segmentations. Adjacent to each measure, the effect sizes of the pairwise comparisons (paired t-tests) between the tools are denoted using Hedges’ g (corrected Cohen’s d), with color-coded indications of magnitude: small (yellow), medium (orange), or large (red). Additionally, the significance levels attained are also presented (* marks p<0.05 and *** marks p<0.001).

#### Interclass correlation coefficient

The analysis of method consistency revealed that all tools had high consistency, with SAMSEG being the tool with the highest ICC (0.99), followed by BIANCA (0.93), LST-LPA (0.91) and LST-LGA (0.88).

#### Sensitivity

LST-LPA showed the highest value (mean: 0.5 ± 0.3), followed by BIANCA (0.4 ± 0.2), LST-LGA (0.4 ± 03) and SAMSEG (0.4 ± 0.3). Statistical analysis revealed significant differences among methods except between BIANCA and SAMSEG (**see Figure 3, top right panel**). The largest sensitivity discrepancies were seen between LST-LPA and the others: BIANCA (D = 1.018), LST-LGA (D = 1.284), and SAMSEG (D = 1.115). Comparisons of BIANCA with LST-LGA and SAMSEG with LST-LGA yielded Cohen’s d values of 0.427 and 0.534, respectively. No significant differences were found between BIANCA and SAMSEG

#### Dice score

LST-LPA showed the highest Dice score (mean: 0.5 ± 0.3), followed by BIANCA (0.5 ± 0.2), LST-LGA (0.3 ± 0.2), and SAMSEG (0.4 ± 0.2). The Dice score of LST-LPA was found to be significantly higher (**see Figure 3, middle right panel**) than the values obtained by BIANCA (D = 0.606), LST-LGA (D = 1.087), and SAMSEG (D = 1.172). Regarding BIANCA, its dice scores showed significantly higher values than the ones obtained with LST-LGA (D = 0.851) and SAMSEG (D = 0.970). Eventually, no significant differences were observed between the dice score of LS-LGA and SAMSEG.

#### Precision

Precision scores were 0.6 ± 0.3 for both BIANCA and LST-LPA, 0.3 ± 0.2 for LST-LGA and 0.3 ± 0.3 for SAMSEG. Significant differences in precision were found between LST-LPA, BIANCA, and the other tools (see **Figure 3, bottom right panel**), but not between LST-LPA and BIANCA themselves. Effect sizes were higher when comparing LST-LPA vs LST-LGA (D = 0.822), LST-LPA vs SAMSEG (D = 1.181), BIANCA vs LST-LGA (D = 0.997), and BIANCA vs SAMSEG (D = 1.562) whereas a modest effect size (D = 0.369) was observed when comparing LST-LGA and SAMSEG.

Finally, FreeSurfer’s performance was inferior to tools based on FLAIR segmentation (see **Supplementary material 7B** for more details).

### 3.3. Performance by total lesion volume

Plotting Dice scores, precision, and sensitivity against the total WMH volume on the gold standard revealed a logarithmic influence of WMH burden on the performance (**Figure 4**). We used R^2^ scores and two quantitative measures to highlight method differences (**Figure 4, left**): 1) the 95% value (in mm^3^) indicates the WMH volume achieving 95% performance; and 2) ∫f(x) represents the area under the fitting curve. FreeSurfer’s results are detailed in **Supplementary material 7C**.

**Figure 4.**
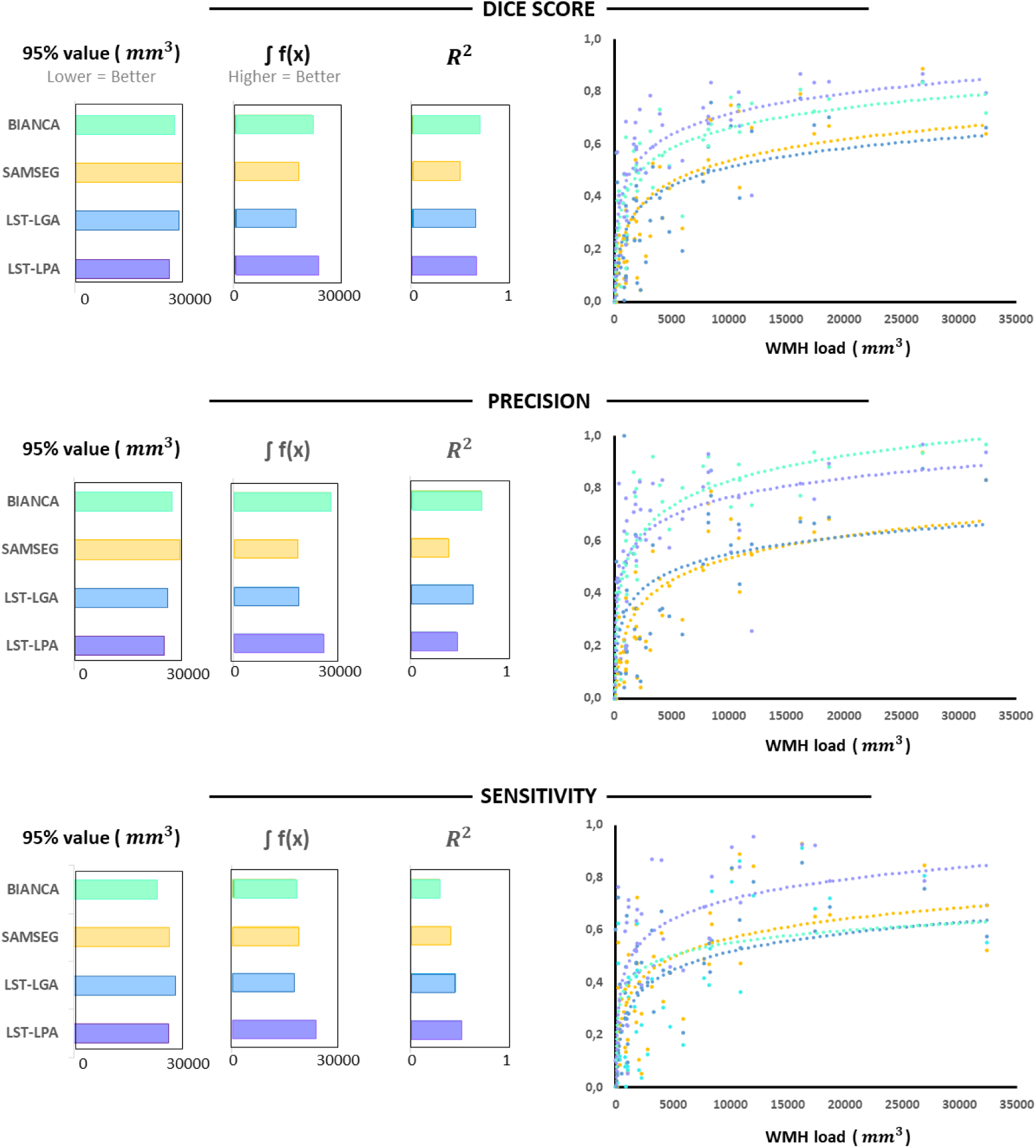
Performance (Dice score, precision, and sensitivity) of each method as a function of the gold standard’s total WMH volume. LST-LPA is depicted in dark blue, LST-LGA in light blue, SAMSEG in yellow, and BIANCA in green. On the right side of the graph, the trend line for each tool and performance measure is shown. On the left, the graph displays the WMH volume that achieves 95% of the maximum performance, the area beneath the curve, and the R^2^ value for each tool and measure. The results for BIANCA are shown in green, SAMSEG in yellow, LST-LGA in light blue, and LST-LPA in dark blue.

For Dice scores, LST-LPA showed a strong logarithmic fit (R2: 0.67), stabilized at a lower WMH volume (95% value: 23230 mm^3^), and had the largest area under the curve (∫f(x) = 26020). BIANCA obtained a R2 of 0.71, 95% value: 24612 mm^3^ and ∫f(x): 24108, whereas LST-LGA (R2: 0.65, 95% value: 25694 mm^3^, ∫f(x): 18870) and SAMSEG (R2: 0.50, 95% value: 26406 mm^3^, ∫f(x): 19840) reached the 95% value at higher volumes and obtained a lower area under the fit curve and a worse log fit.

In precision, BIANCA obtained the best logarithmic fit with a R2 of 0.73, 95% value: 24327 mm^3^ and ∫f(x): 30180. In contrast, LST-LPA (R2: 0.48, 95% value: 22433 mm^3^, ∫f(x): 27733) and LST-LGA (R2: 0.65, 95% value: 23191 mm^3^, ∫f(x): 20092) had poorer log fit and lower area under the fit curve but stabilized at a lower WMH volume. On the other hand, SAMSEG (R2: 0.39, 95% value: 26111 mm^3^, ∫f(x): 19751) was the tool with the worst logarithmic fit, the lowest area under the fit curve and which stabilized at higher values.

For sensitivity, LST-LPA reached an R2 of 0.53, 95% value: 23371 mm^3^ and ∫f(x): 26061, followed by BIANCA (R2: 0.3, 95% value: 20678 mm^3^, ∫f(x): 19871) that stabilized at lower WMH volumes but obtained lower log fit and area under the fitting curve. By contrast, SAMSEG (R2: 0.42, 95% value: 23694 mm^3^, ∫f(x): 20849), and LST-LGA (R2: 0.47, 95% value: 25086 mm^3^, ∫f(x): 18981) had lower R2, ∫f(x) and stabilized at higher WMH values.

### 3.4. Performance by individual lesion volume

To assess the sensitivity of each tool in detecting individual lesions as defined by the gold standard, we stratified our analysis by lesion volume. **Figure 5** illustrates the sensitivity of each tool across different lesion volume ranges. The initial finding was that the majority of lesions 8 mm^3 or smaller were not detected by any of the tools. Moreover, the trend of sensitivity relative to lesion size exhibited distinct patterns for each tool. Additionally, we investigated whether any tool significantly surpassed a sensitivity threshold of 0.5 for any lesion group. The outcome was that only the LST-LPA algorithm demonstrated a sensitivity higher than 0.5, specifically within the lesion group larger than 111 mm^3. FreeSurfer results are presented in **Supplementary material 7D**.

**Figure 5.**
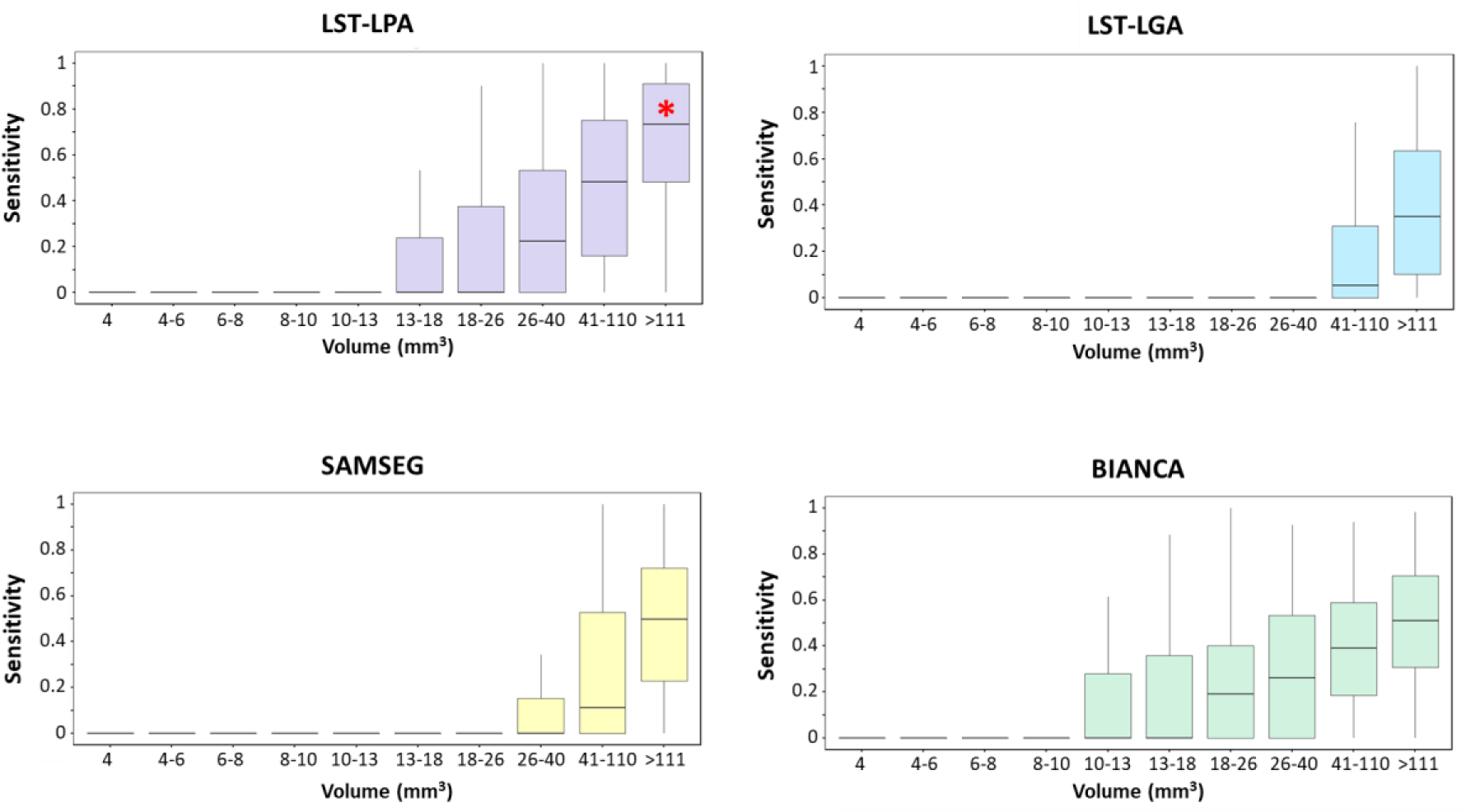
The sensitivity of each tool when assessing individual lesions stratified by volume ranges is presented. Each volume range includes an equal number of lesions (196). For every tool and volume range, the boxplot displays the median, as well as the second and third quartiles. The whiskers extend to the first and fourth quartiles, excluding outlier individuals. The asterisk (*) denotes groups that achieved a sensitivity significantly greater than 0.5.

### 3.5. Real world scenario and outlier evaluation

Based on the results obtained, we decided to test the consistency of the differences in a sample of 300 elderly individuals from our research group using both LST algorithms. The LST tools were chosen for their performance, particularly the LST-LPA, and for their rapid processing and user-friendliness. The findings indicated that LST-LPA experienced malfunctions in 2 patients, whereas LST-LGA had issues in 3 patients. Notably, the results suggested that using both LST tools together provided a reliable estimate of WMH volumes, with only those cases showing large discrepancies between the two tools’ results requiring visual verification. **Figure 6** illustrates the differences in the outputs of these tools for the sampled individuals and includes MRI images for cases in which WMH segmentation was suboptimal.

**Figure 6.**
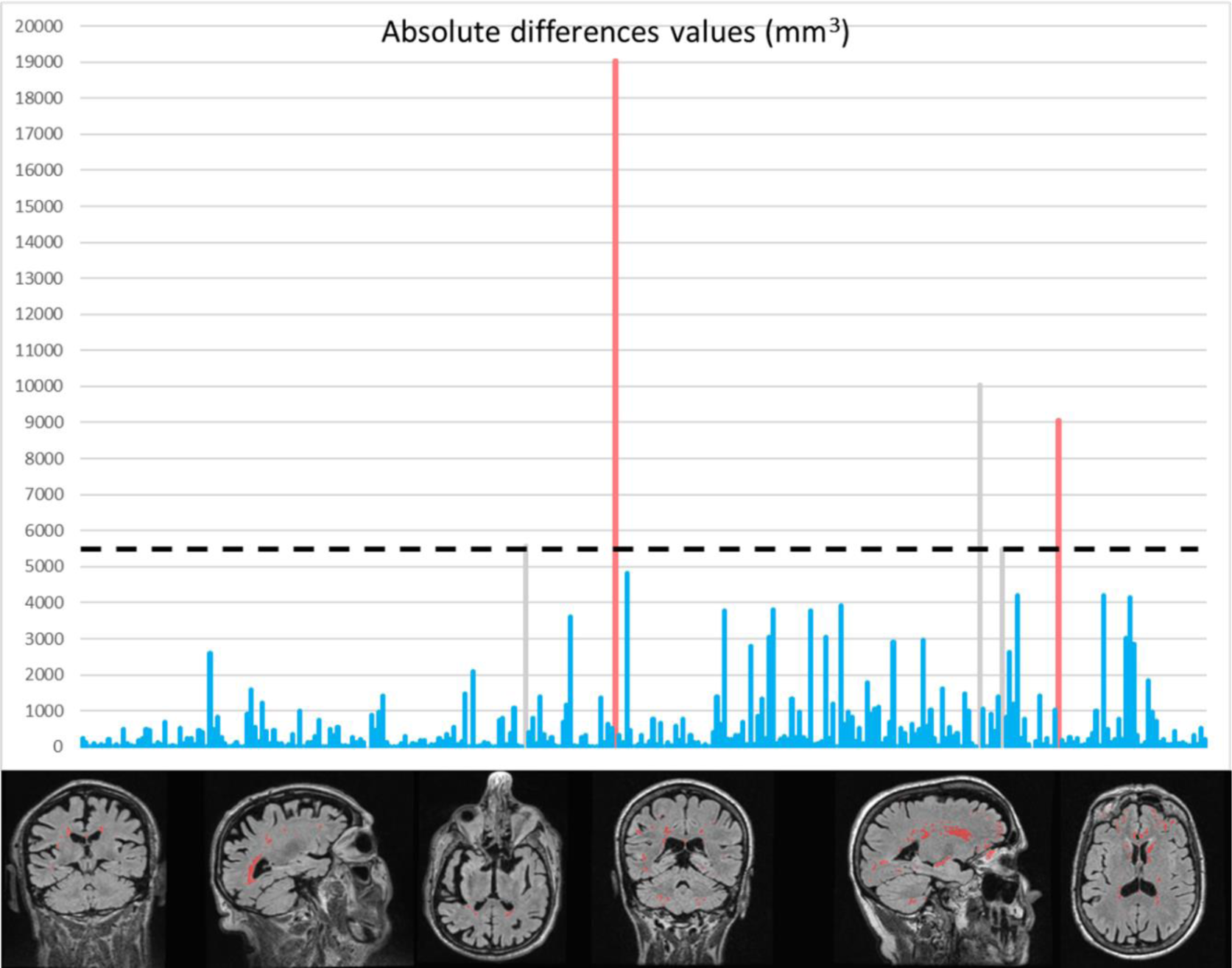
Representation of absolute differences between LST-LPA and LST-LGA across 300 subjects. The dashed line indicates the outlier threshold, set at three standard deviations above the mean. Blue bars represent subjects whose differences fall below the outlier threshold and who have good quality segmentations according to the LST-LPA tool. Grey bars identify outliers with good WMH segmentation as determined by the LST-LPA tool. Red bars highlight outliers with poor WMH segmentation by the LST-LPA tool. Examples of suboptimal LST-LPA segmentations are provided at the bottom of the figure.

### 3.6. Biomarker extraction and sample characterization

The obtained cut point corresponded to a WMH volume of 960 mm^3^ (¡Error! No se encuentra el origen de la referencia.), which, for the purposes of characterizing the complete sample, was rounded to 1000 mm^3^. Based on this cut-off, we divided the sample into two subgroups: those with a low vascular load (Low-VL), defined as having less than 1000 mm^3^ of WMH, and those with a mild vascular load (Mild-VL), defined as having WMH volumes above this threshold (see **Figure 7**). It was observed that 59% of the population fell into the Low-VL category, while the remaining 41% had WMH volumes exceeding the cut point. The latter group demonstrated an exponential increase in WMH volume, reaching up to 23203 mm^3^.

**Figure 7.**
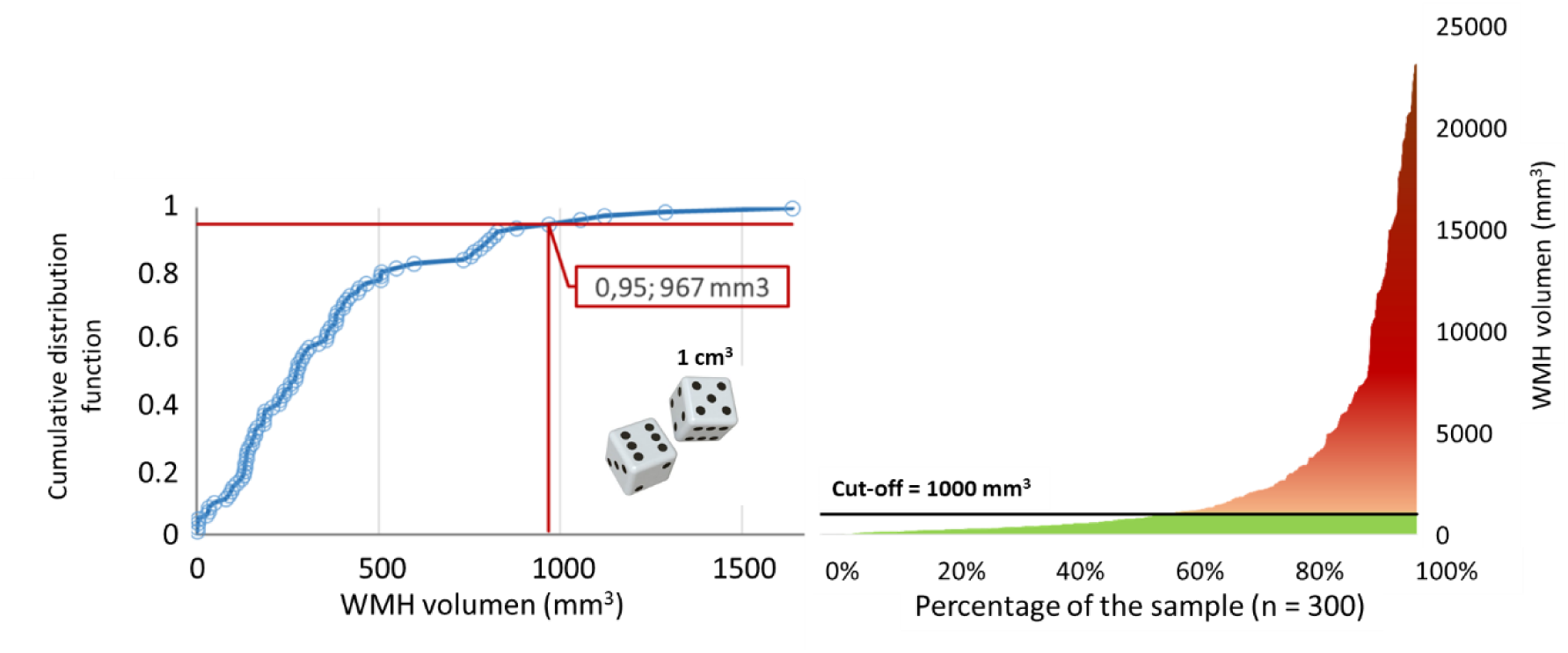
On the left, a graphical summary illustrates the method employed to determine the WMH volume cut-off point. The cumulative distribution function is plotted with WMH volume on the horizontal axis and the proportion of observations with WMH volume less than or equal to that on the vertical axis. This is visually represented by a dice with a side length of 1 cm, the volume of which corresponds to the cut-off point for WMH volume. On the right, the total sample distribution is categorized based on cerebrovascular damage, as indicated by WMH volume.

To characterize the populations divided according to the cut-off point, we investigated potential sociodemographic, behavioral, and brain integrity differences associated with the severity of vascular damage, with the findings detailed in **Table 2**. For clarity, we have organized the results into four categories: sociodemographic, cognitive performance, brain structural integrity, and white matter integrity. Sociodemographically, age was the sole significant difference between the WMH volume-based subgroups. Cognitively, only delayed recall showed a significant difference (p=0.041). No notable differences were found in standard dementia markers like gray matter or hippocampal volume. Yet, significant differences were observed in SVD-related neuroimaging markers, specifically lateral ventricle enlargement (p=1E-08) and reduced white matter volume (p=0.039). Differences in white matter integrity were noted in certain projection fibers, thalamic radiations, and association fibers, but post-FDR correction, only lateral ventricle and white matter integrity differences remained significant. See **Table 2** for detailed p-values.

**Table 2.** Sample characterization across all collected variables. Values are presented as mean ± SD. MMSE = Mini-Mental State Examination, TMT-A = Trail-Making Test part A, TMT-B = Trail-Making Test part B, GM = grey matter, WM = white matter. p-values and F scores correspond with between-groups ANCOVA test with age and total brain volume segmentation as covariates. Symbol * depict those p-values that survived the FDR correction. For age and brain segmentation volume we used between-groups ANCOVA test excluding the correspondent as covariate. For sex and ApoE-ε4 we used Fisher exact test. η2 account for partial-Eta squared for all comparisons but for the case of age and brain segmentation, where the values correspond to η2 values.

| | | Mild VI (124) | Low VI (176) | p-value | F value | Partial $\eta^2$ |
| --- | --- | --- | --- | --- | --- | --- |
| Sample features | Sex (females) | 63% | 71% | 0,168 |  |  |
| | ApoE-ε4 | 30% | 27% | 0,683 | | |
| | Age | 72 ± 6 | 62 ± 7 | 9E-27* | 140,35 | 0,321 |
| | Education (years) | 14 ± 5 | 14 ± 5 | 0,533 | 0,39 | 0,002 |
| | MMSE | 29 ± 1 | 29 ± 1 | 0,330 | 0,95 | 0,003 |
| Cognitive performance | Immediate Recall | 35 ± 12 | 39 ± 11 | 0,106 | 2,63 | 0,009 |
| | Delayed Recall | 21 ± 10 | 25 ± 8 | 0,041 | 4,23 | 0,015 |
| | Digit Spam Forward | 9 ± 2 | 9 ± 2 | 0,286 | 1,14 | 0,004 |
| | Digit Spam Backward | 6 ± 2 | 6 ± 2 | 0,462 | 0,54 | 0,002 |
| | Phonemic Fluency | 14 ± 5 | 14 ± 4 | 0,852 | 0,03 | 1E-04 |
| | Semantic Fluency | 17 ± 3 | 19 ± 4 | 0,212 | 1,57 | 0,006 |
| | TMT A (time) | 52 ± 20 | 44 ± 18 | 0,892 | 0,02 | 7E-05 |
| | TMT B (time) | 133 ± 75 | 96 ± 56 | 0,132 | 2,28 | 0,008 |
| Brain structure (Volume in mm<sup>3</sup>) | Brain segmentation | 1042299 ± 114543 | 1062790 ± 103158 | 0,069 | 3,32 | 0,011 |
| | GM | 0,53 ± 0,02 | 0,54 ± 0,03 | 0,214 | 1,55 | 0,005 |
| | Cortical GM | 0,39 ± 0,02 | 0,39 ± 0,02 | 0,386 | 0,75 | 0,003 |
| | Subcortical GM | 0,047 ± 0,003 | 0,047 ± 0,002 | 0,792 | 0,07 | 2E-04 |
| | WM | 0,40 ± 0,02 | 0,42 ± 0,03 | 0,039 | 4,29 | 0,014 |
| | Cortical thickness (mm) | 2,3 ± 0,1 | 2,3 ± 0,1 | 0,220 | 1,51 | 0,005 |
| | Lateral ventricles | 0,02 ± 0,01 | 0,01 ± 0,005 | 1E-08* | 34,25 | 0,104 |
| | Hippocampus | 0,007 ± 0,001 | 0,007 ± 0,001 | 0,146 | 2,12 | 0,007 |
| WM microstructure (Fractional Anisotropy) | Forceps Minor (fmi) | 0,39 ± 0,02 | 0,40 ± 0,02 | 0,003* | 9,08 | 0,032 |
| | Forceps Major (fma) | 0,39 ± 0,02 | 0,40 ± 0,02 | 0,003* | 8,89 | 0,032 |
| | Middle cerebellar peduncle (mcp) | 0,50 ± 0,04 | 0,50 ± 0,03 | 0,734 | 0,12 | 4E-04 |
| | Medial lemniscus (ml) | 0,44 ± 0,02 | 0,46 ± 0,02 | 7E-07* | 25,89 | 0,087 |
| | Corticospinal tract (cst) | 0,41 ± 0,02 | 0,42 ± 0,02 | 0,094 | 2,83 | 0,010 |
| | Acoustic radiation (ar) | 0,54 ± 0,02 | 0,54 ± 0,02 | 0,123 | 2,40 | 0,009 |
| | Anterior thalamic radiation (atr) | 0,47 ± 0,02 | 0,19 ± 0,02 | 6E-06* | 21,33 | 0,073 |
| | Superior thalamic radiation (str) | 0,43 ± 0,02 | 0,44 ± 0,02 | 0,002* | 10,17 | 0,04 |
| | Posterior thalamic radiation (ptr) | 0,40 ± 0,02 | 0,41 ± 0,02 | 0,179 | 1,82 | 0,007 |
| | Superior longitudinal fasciculus (slf) | 0,45 ± 0,03 | 0,45 ± 0,02 | 0,759 | 0,09 | 3E-04 |
| | Inferior longitudinal fasciculus (ilf) | 0,50 ± 0,02 | 0,47 ± 0,08 | 0,002* | 9,98 | 0,036 |
| | Inferior fronto-occipital fasciculus (ifo) | 0,44 ± 0,03 | 0,44 ± 0,03 | 0,255 | 1,30 | 0,005 |
| | Uncinate fasciculus (unc) | 0,47 ± 0,02 | 0,47 ± 0,02 | 0,884 | 0,02 | 8E-05 |
| | Cingulate gyrus part of cingulum (cgc) | 0,51 ± 0,03 | 0,53 ± 0,02 | 1E-08* | 34,21 | 0,112 |
| | Parahippocampal part of cingulum (cgh) | 0,41 ± 0,03 | 0,41 ± 0,02 | 0,911 | 0,01 | 5E-05 |

## 4 Discussion

In this study, we aimed to guide clinicians and researchers in choosing the most suitable method for evaluating and monitoring white matter hyperintensities (WMH) of suspected vascular origin. We assessed the performance of four open-access algorithms (LST-LPA, LST-LGA, SAMSEG, and BIANCA) for WMH segmentation in MRI across a sample of 45 cognitively healthy older adults with varying WMH loads. These algorithms are part of three popular neuroimaging toolboxes (FSL, FreeSurfer, and SPM). We evaluated each tool by correlating its output with the Fazekas scale and radiologist manual segmentations, considering lesion-level sensitivity. The algorithms generally correlated well with clinical scales and the manual gold standard, with LST-LPA and BIANCA performing best. However, we chose to focus further on the LST tools for their complementary strengths. The two LST algorithms were applied to a larger dataset of 300 seniors to test consistency and defined a vascular damage biomarker at the 95th percentile of WMH volume. Using the established cut-off, we identified two distinct groups with significant differences in brain integrity metrics and potential cognitive decline trends.

### 4.1 Where the automatic results related to the clinical status?

The automated tools demonstrated strong correlations with Fazekas scores, nearly matching the gold standard in total WMH volume estimation, despite a tendency to underestimate lesion volumes (**see Supplementary material 5 & 6**). These findings underscore the clinical utility of automated segmentation as a reliable surrogate for the Fazekas scale and manual segmentation. By combining Fazekas scores with total WMH volume, we gain a robust measure of vascular damage, balancing the Fazekas scale’s limitations with the granularity of WMH volume assessment. Previous studies have validated the correlation between automatic and manual segmentations for LST-LPA [40], [41], LST-LGA [40], [41], and BIANCA [31], [42], with literature indicating a general trend for these tools to report smaller lesion volumes compared to manual segmentation [31], [41]. However, no previous SAMSEG’s correlation with manual segmentation were found in the literature. As can be seen in **Supplementary material 6**, LST-LPA showed the highest concordance with manual segmentation, with LST-LGA and SAMSEG also performing well. These findings diverge from those reported by [40], where LST-LGA outperformed LST-LPA. In contrast, BIANCA was less aligned with clinical measurements but fitted regression models best, suggesting a potential systematic issue in handling false negatives, as indicated by sensitivity analyses (**Figures 3 and 4**).

### 4.2 General performance comparison: total volume matters

All algorithms displayed high stability, with SAMSEG achieving the highest ICC, followed by BIANCA and the LST tools. While previous studies on LST-LPA, LST-LGA, and BIANCA report mixed ICC results, our findings are in line with some and differ from others (Griffanti et al., 2016; Heinen et al., 2019; Hotz et al., 2021; Tran et al., 2022). We assessed the correlation between automated WMH segmentation and manual radiologist segmentation by analyzing precision, sensitivity, and Dice scores. LST-LPA was identified as the most sensitive and closely matched algorithm to manual segmentation. Regarding precision, LST-LPA and BIANCA obtained comparable values, surpassing SAMSEG and LST-LGA (**Figure 3**). Consequently, our results pointed out that supervised methods (LST-LPA and BIANCA), which use complex algorithms such as logistic regression [43] and K-nearest neighbors [42], respectively, outperformed the unsupervised methods. Algorithm performance, in terms of sensitivity, accuracy, and Dice score, increased logarithmically with higher lesion burden (**Figure 4**), confirming previous findings [30], [31] that effectiveness diminishes in cases with minimal vascular damage. Our results highlighted that LST-LPA showed the most significant improvement in Dice score and sensitivity with increasing WMH volume compared to BIANCA. LST-LPA also reached the cut-off value faster than BIANCA in both precision and Dice score, although BIANCA exhibited a quicker improvement in sensitivity. Among the unsupervised methods, SAMSEG demonstrated superior performance in sensitivity, whereas LST-LGA excelled in precision.

Our results align with prior findings on supervised tools [40], [44] and SAMSEG [45], yet LST-LGA underperformed in our analysis [40], [46]. Similar patterns of algorithm performance varying with lesion volume have been noted, with other studies reporting more effective results at lower levels of vascular damage [41], [42], [47]. The performance disparity might stem from the use of MRI scanners with different magnetic field strengths, as LST-LGA’s accuracy is compromised on the 1.5 tesla machines commonly used in clinical settings due to their lower resolution, especially for small lesions [45]. This is particularly relevant for LST-LGA, which depends on T1 hypointensities to initialize lesions that will subsequently grow on the FLAIR image [47]. As mentioned by the authors of LST-LGA, hypointensities in T1 images are sensitive to field strength, and may not exist for certain lesions on low-resolution scans [47].Moreover, lesion etiology [25] could play a role, as most comparative studies focused on multiple sclerosis patients.

### 4.3 Individual lesion size bias

Detecting small lesions is crucial in clinical and scientific research, particularly in studies on healthy aging or early stages of pathological aging, where vascular damage is often minimal, and in longitudinal studies tracking disease progression. The location and distribution patterns of WMH are linked to various pathologies [25], [48] and cognitive impairments [49]. While automatic segmentation tools are generally validated for their sensitivity, precision, and Dice scores, their performance on smaller lesions is less impactful on these global measures due to differences in lesion volume, definition, and etiology [25], [50]. Moreover, metrics sensitive to the surface-volume ratio of lesions require careful consideration of lesion size for accurate performance evaluation [30], [32]. Small lesions present challenges due to their intensity range affected by the partial volume effect at tissue boundaries, leading to non-homogeneity [25], [32], [50]–[52]. Additionally, their blob-like structures and variable locations contrast with larger lesions typically found around the periventricular area [50]. This heterogeneity makes small lesion segmentation a complex, nonlinear problem for automatic algorithms, impacting their performance [30]. We addressed this by measuring the sensitivity for each lesion individually.

Our findings indicate that lesion size significantly impacts segmentation quality, with each tool showing variable performance based on lesion volume (**see Figure 5**). BIANCA maintains consistent sensitivity for lesions from 10 to 13 mm³, aligning with previous research [51]. LST-LPA achieves similar consistency at slightly larger sizes, between 13 and 18 mm³. Both tools show a marked sensitivity increase for lesions over 26 mm³, but only LST-LPA exceeds a sensitivity of 0.5 for larger lesions. Unsupervised methods only reliably segment lesions above 26 mm³, with SAMSEG being marginally more sensitive. This limitation may stem from difficulties in delineating lesion borders, particularly for smaller lesions with high surface-to-volume ratios. In summary, although BIANCA and LST-LPA perform well with small lesions, they fail to consistently segment those smaller than 8 mm³. Moreover, no algorithm consistently achieves over 50% sensitivity for lesions larger than 111 mm³, leading to an underestimation of total vascular burden. Recent machine learning approaches show promise for small lesion segmentation, utilizing multiscale deep features [51] or separate systems for small and large lesions [50]. However, these methods need external validation and testing on larger and more diverse samples to confirm their effectiveness.

### 4.4 Comparison between algorithms: Which algorithm should be used?

Supervised methods outperform unsupervised ones in lesion segmentation, achieving higher sensitivity and precision, partly due to their ability to use more diverse characteristics to identify lesions [32]. LST-LPA surpasses BIANCA in sensitivity and Dice score, offering high-quality segmentation and better performance with larger lesions. Although LST-LPA shows less consistency with small lesions, it effectively segments them and demonstrates increased sensitivity with larger lesions. Methodologically, the primary distinction between the two algorithms lies in their preparation; BIANCA is trained on a sample akin to the gold standard, whereas LST-LPA employs a pre-trained algorithm on multiple sclerosis patient samples. The original authors have indicated that they are developing a version of LST-LPA that can tailor the learning process to individual samples (Schmidt, 2016). In terms of preprocessing, BIANCA often encounters issues when setting exclusion masks, resulting in numerous false positives—a challenge in managing large databases. Given these considerations, along with its user-friendliness, minimal computational demands, and quick processing times, LST-LPA stands out as one of the superior options available. Attending to unsupervised methods, LST-LGA has higher precision, but lower sensitivity compared to SAMSEG, with the latter detecting smaller lesions. LST-LGA stands out with its user-friendly interface, minimal computational load, and rapid processing. In contrast, SAMSEG offers comprehensive data, including volumetrics of cortical and subcortical areas, valuable for academic research.

Considering other algorithms not evaluated in this work, recent advancements have seen convolutional neural networks (CNNs) outperforming traditional methods like LST-LPA in some cases, unaffected by vascular load [44]. Additionally, methods considering spatial intensity variability have been proposed to enhance segmentation accuracy [53]. Despite these innovations, CNN-based tools have not consistently outperformed simpler methods such as LST-LPA and BIANCA. This suggests that while newer techniques offer potential improvements, traditional tools remain competitive due to their specificity [25].

Considering our results, we employed both LST tools for WMH segmentation in a larger older adult cohort due to their optimal quality-cost ratio, sharing a toolbox with low computational demands and swift processing. Among 300 participants, we identified 5 outliers based on discrepancies in WMH volume between the tools. Visual checks (**Figure 6, bottom panel**) revealed that LST-LPA inaccurately marked false positives in two cases, while LST-LGA failed in three. One possible explanation is that the advantage of LST-LGA in some of these segmentations may be due to its independence from manual segmentations of varying reliability [31], [40]. Studies show that while complex algorithms like CNNs struggle with routine clinical images, simpler methods like LST-LPA and BIANCA are more robust [46]. Despite this, managing large datasets with these tools still demands significant review and quality control. However, our findings suggest that the combined use of both LST tools provides a reliable, efficient approach for WMH segmentation.

### 4.5 Clinical relevance: How automated tools can define early biomarkers of vascular damage?

Due to the impracticality of manual segmentation by radiologists for our entire database, we explored the efficacy of automatic segmentation tools for assessing vascular damage. Accurate WMH assessment is crucial as it influences diagnosis, particularly with low lesion volumes. LST-LPA closely matched the gold standard, adapting quickly with vascular load, which led to its selection for subsequent analysis. Our findings revealed in our sample a significant WMH volume variability, suggesting an exponential increase beyond a certain threshold. Investigating a potential clinical tipping point, we established a WMH volume cutoff at 1000 mm³ for the healthiest adults aged 50-60, following the guidelines by Jack et al. (2017).

Previous studies suggest that WMH progression is non-linear and dynamic, with age accounting for roughly a third of WMH volume variance [54], [55]. Our cross-sectional study, spanning ages 50 to 84, confirms age’s influence on WMH quantity, with notable WMH total volume differences across age-defined groups (**Table 2**). Although vascular damage is linked to cognitive decline [8], [9], our sample, divided by vascular burden, showed no significant cognitive differences post-FDR correction, indicating early-stage disease with preserved cognition. However, structural brain changes were evident, including ventricular dilatation and altered white matter tract integrity, particularly in connections from deep brain regions [17], [18]. No gray matter deterioration or ApoE ε4 allele-related differences emerged, likely due to our healthy aging cohort. In essence, a vascular damage biomarker distinguished groups with structural brain differences and potential cognitive decline risk, underscoring its value in identifying high-risk individuals in clinical and research settings.

### 4.6 Limitations and Future research

Despite the study’s contributions, we must acknowledge its methodological limitations. The reliance on manual segmentation as a gold standard is problematic due to the subjective nature of WMH definitions and the resulting inter-rater variability [32], [46]. Moreover, using 1.5 tesla MRI scanners, while clinically relevant, hinders comparison with studies employing higher-resolution 3 tesla MRI images. These factors collectively challenge comparisons with prior studies and could account for some observed discrepancies. The proliferation of automatic segmentation methods presents an opportunity for advancement, yet the lack of access to these tools or their proprietary algorithms complicates efforts to compare and improve upon them. To address this, our work focused solely on publicly available tools with established use in clinical and research settings. A standardized consensus on WMH segmentation is critical for combining different methodologies and enhancing precision. Furthermore, due to variations in sample characteristics and technical specifications across studies, WMH measurements may not be directly comparable [55]. Therefore, the cutoff value proposed in this thesis is a preliminary benchmark specific to our sample and requires further validation before it can be considered clinically generalizable.

## 5 Conclusion

Selecting the right WMH segmentation method depends on the desired metrics and sample traits. For assessing total WMH volume, unsupervised tools reliably mirror manual segmentation. In contrast, when identifying lesion locations or handling populated samples of small lesions, supervised methods excel. However, the importance of quality control cannot be underestimated; it’s essential for detecting segmentation anomalies. In our experience, the LST suite stands out, offering two robust, easy-to-use options: the supervised LST-LPA and the unsupervised LST-LGA. These can be employed synergistically for reliable WMH segmentation The utilization of these methods has led to the accurate determination of WMH volumes on MRI scans, ensuring a reliable index of vascular damage and improving the consistency of research findings. Furthermore, they have been instrumental in establishing a biomarker threshold for vascular damage, associated with structural brain alterations and a potential cognitive decline, anchoring these measures in clinical significance.

## 6 Founding

Lucia Torres-Simon and Alberto del Cerro-León acknowledge the financial support of predoctoral researchers grants from Universidad Complutense de Madrid (CT42/18-CT43/18 & CT58/21-CT59/21, respectively), that were co-funded by Santander bank. Database was supported by three Spanish projects: PSI2009-14415-C03-01, PSI2012-38375-C03-01 and PSI2015-68793-C3-1-R.

Research reported in this publication was supported partially (FM, PC) by the National Institute on Aging of the National Institutes of Health under award numbers RF1AG074204 and RF1AG079324. The content is solely the responsibility of the authors and does not necessarily represent the official views of the National Institutes of Health.

## 8 Compliance with ethical standards

The Hospital Universitario San Carlos Ethics Committee (Madrid) approved the study, and all participants signed a written informed consent prior to participation.

